# 30-day mortality in patients hospitalized with COVID-19 during the first wave of the Italian epidemic: a prospective cohort study

**DOI:** 10.1101/2020.05.02.20088336

**Authors:** Andrea Giacomelli, Anna Lisa Ridolfo, Laura Milazzo, Letizia Oreni, Dario Bernacchia, Matteo Siano, Cecilia Bonazzetti, Alice Covizzi, Marco Schiuma, Matteo Passerini, Marco Piscaglia, Massimo Coen, Guido Gubertini, Giuliano Rizzardini, Chiara Cogliati, Anna Maria Brambilla, Riccardo Colombo, Antonio Castelli, Roberto Rech, Agostino Riva, Alessandro Torre, Luca Meroni, Stefano Rusconi, Spinello Antinori, Massimo Galli

## Abstract

**Background:** Italy was the first European country hit by the COVID-19 pandemic and has the highest number of recorded COVID-19 deaths in Europe.

**Methods:** This prospective cohort study of the correlates of the risk of death in COVID-19 patients was conducted at the Infectious Diseases and Intensive Care units of Luigi Sacco Hospital, Milan, Italy. The clinical characteristics of all the COVID-19 patients hospitalised in the early days of the epidemic (21 February -19 March 2020) were recorded upon admission, and the time-dependent probability of death was evaluated using the Kaplan-Meier method (censored as of 20 April 2020). Cox proportional hazard models were used to assess the factors independently associated with the risk of death.

**Results:** Forty-eight (20.6%) of the 233 patients followed up for a median of 40 days (interquartile range 33-47) died during the follow-up. Most were males (69.1%) and their median age was 61 years (IQR 50-72). The time-dependent probability of death was 19.7% (95% CI 14.6-24.9%) 30 days after hospital admission. Age (adjusted hazard ratio [aHR] 2.08, 95% CI 1.48-2.92 per ten years more) and obesity (aHR 3.04, 95% CI 1.42-6.49) were independently associated with an increased risk of death, which was also associated with critical disease (aHR 8.26, 95% CI 1.41-48.29), C-reactive protein levels (aHR 1.17, 95% CI 1.02-1.35 per 50 mg/L more) and creatinine kinase levels above 185 U/L (aHR 2.58, 95% CI 1.37-4.87) upon admission.

**Conclusions:** Case-fatality rate of patients hospitalized with COVID-19 in the early days of the Italian epidemic was about 20%. Our study adds evidence to the notion that older age, obesity and more advanced illness are factors associated to an increased risk of death among patients hospitalized with COVID-19.

## BACKGROUND

In late December 2019, an outbreak of an emerging disease (COVID-19) caused by a novel coronavirus that was later named severe acute respiratory syndrome coronavirus 2 (SARS-CoV-2) was recorded in Wuhan, China [1], and subsequently rapidly spread to a substantial number of Asian and non-Asian countries. It was declared a pandemic by the World Health Organisation on 12 March 2020 [2].

The increasing number of studies conducted in Chinese hospitals over the last few months have contributed to delineating the characteristics of the disease and its lethality [3–6]. They describe COVID-19 as an atypical SARS-like pneumonia that requires intensive care in 26-33% of patients, 4-15% of whom eventually die [4, 5]. However, as the epidemic moves outside China, there is a need to verify the clinical features and lethality of COVID-19 in countries with different demographic characteristics.

Italy was the first European country to be hit hard by the COVID-19 epidemic, with Lombardy in northern Italy being the region in which the first autochthonous cases were identified and the largest epidemic foci developed [7]. Italy is also the European country in which the highest number of COVID-19 deaths have so far been recorded (24,780 as of 27 April 2020) [8].

The Department of Infectious Diseases of Luigi Sacco Hospital (the national reference centre for epidemiological emergencies and bioterrorism in northern Italy) has been admitting SARS-CoV-2 patients (particularly those coming from the “red zone” municipalities first involved in the epidemic) since the night of 20 February 2020, when the first COVID-19 case was identified in a town about 50 km from Milan [9].

This paper describes the demographic and clinical characteristics of the COVID-19 patients admitted to our hospital between 21 February and 19 March 2020 in the early stage of the Italian epidemic, and the factors associated with the risk of COVID-19-related death.

## METHODS

### Setting

The study was conducted in the continuously evolving scenario created by the dramatic escalation of the COVID-19 epidemic in Lombardy. The large structural changes that had to be made in the organisation of our hospital over a 2-week period transformed our 68-bed Department of Infectious Diseases and 8-bed general intensive care unit (ICU) into a single-building isolation area with 93 non-intensive and 30 intensive care beds entirely dedicated to COVID-19 patients.

### Study design and participants

This was a single-centre, prospective cohort study of all of the adult COVID-19 patients admitted to Luigi Sacco Hospital in Milan, Italy, between 21 February (the day the first patients were hospitalised) to 19 March 2020; the observation of the cohort was censored on 20 April 2020. All of the study patients had COVID-19 confirmed by a positive real-time reverse-transcription polymerase chain reaction on a nasopharyngeal swab.

### Data collection

The data extracted from the patients’ clinical charts on a daily basis and stored in an *ad hoc* database included age and gender; the reported date of symptom onset; body weight and height, the presence of obesity defined as a body mass index ≥ 30 points [10], and history of smoking; comorbidity burden defined assessed by age unadjusted Charlson comorbidity index [11] and concomitant treatments for chronic medical conditions; symptoms; vital signs (heart rate, respiratory rate, blood pressure, pulse oximetry), laboratory values (white blood cell, neutrophil, lymphocyte, and platelet counts; hemoglobin, albumin, lactate dehydrogenase, C-reactive protein (CRP), creatine kinase (CK), alanine aminotransferase, bilirubin, prothrombin, D-dimer, and creatinine levels; and arterial oxygen partial pressure); radiography findings upon admission. The chest X-ray images were reviewed and categorised as follows: no pathological findings; interstitial changes; monolateral lung consolidation (s); bilateral lung consolidation(s); and pleural effusion (yes/no). Oxygen therapy support started upon hospital admission, and its type (simple face mask, face mask with oxygen reservoir bag, Venturi-type oxygen mask, continuous positive airway pressure device (cPAP), and mechanical ventilation) were collected.

Using the criteria proposed by Wu *et al*. [12], disease severity upon admission was classified as mild (mild clinical symptoms, no imaging signs of pneumonia); moderate (fever, cough, dyspnoea or other symptoms, imaging signs of pneumonia); severe (any of: respiratory distress with a respiratory rate (RR) of ≥ 30 breaths per minute; resting oxygen saturation in air ≤93%; or PaO_2_ / FiO_2_ ≤ 300 mmHg); and critical (any of respiratory failure requiring mechanical ventilation; shock; or any other organ failure needing intensive care).

Data on the use of antivirals [lopinavir/ritonavir (LPV/r) and remdesivir], and/or antibiotic and/or immunomodulatory agents [hydroxychloroquine (HCQ), tocilizumab] during hospitalization were also collected.

The primary outcome of interest was death; the life status of the patients discharged before the censoring date was ascertained by means of telephone calls made by two physicians on 20 April, 2020.

### Data analysis

The descriptive statistics include proportions for categorical variable, and median values and interquartile range (IQR) for continuous variables. The baseline demographic and clinico-epidemiological characteristics of the survivors and non-survivors were compared using *χ*^2^ or Fisher’s exact test where necessary for categorical variables and Wilcoxon’s rank-sum test for continuous variables.

The time-dependent probability of death during the study period was assessed using the Kaplan-Meir method.

The association(s) between clinically relevant, non-collinear and complete variables (without any missing data upon hospital admission) and the primary outcome was assessed by means of uni- and multivariable Cox proportional hazard models. The multivariable analysis was made by introducing into the model the variables that found to be significantly associated with outcome in the univariate analysis, as well as potential confounders.

All of the statistical analyses were made using SAS software, version 9.4, and differences with *P* values of <0.05 were considered statistically significant.

The study was approved by our Comitato Etico Interaziendale Area 1. Informed consent was waived in the case of patients undergoing mechanical ventilation upon admission.

## RESULTS

Between 21 February and 19 March 2020, a total of 233 COVID-19 patients were admitted to L. Sacco Hospital, Milan, Italy. Most were males (69.1%) of Italian nationality (92.8%), and their median age was 61 years (IQR 50-72). Twenty-six (11.2%) were healthcare workers. A total of 133 (57.1%) were resident in the city or metropolitan area of Milan (the first case in Milan was recorded on 23 February 2020). Twenty-seven (11.6%) came from the “red zones” and 71 were transferred from other provinces in Lombardy (Lodi, Cremona, and Bergamo) whose hospitals were overwhelmed by the sudden explosion of the epidemic.

Forty-eight patients (20.6%) died during the study period, and 185 survived, including 162 (69.5%) patients who were discharged, and 23 (9.9%) who were still hospitalised on the censoring date.

Table 1 shows the differences in the baseline demographic and clinical characteristics of the survivors and non-survivors. The non-survivors included a higher proportion of subjects aged 66-75 (39.6% *vs* 19.5%), 76-85 (20.8% *vs* 13.0%), and 86-95 years (10.4% *vs* 2.7%) (p<0.001); a higher proportion of patients transferred from other hospitals (62.5% *vs* 36.8%, p<0.002). The non-survivors were more frequently being treated with anti-platelet agents (p=0.009), calcium channel blockers (p=0.023) and angiotensin II receptor blockers (p=0.001). Conversely, the survivors were more frequently without any co-medication on hospital admission when compared to the non-survivors (37.8% *vs* 18.8%, p=0.016).

**Table 1.**
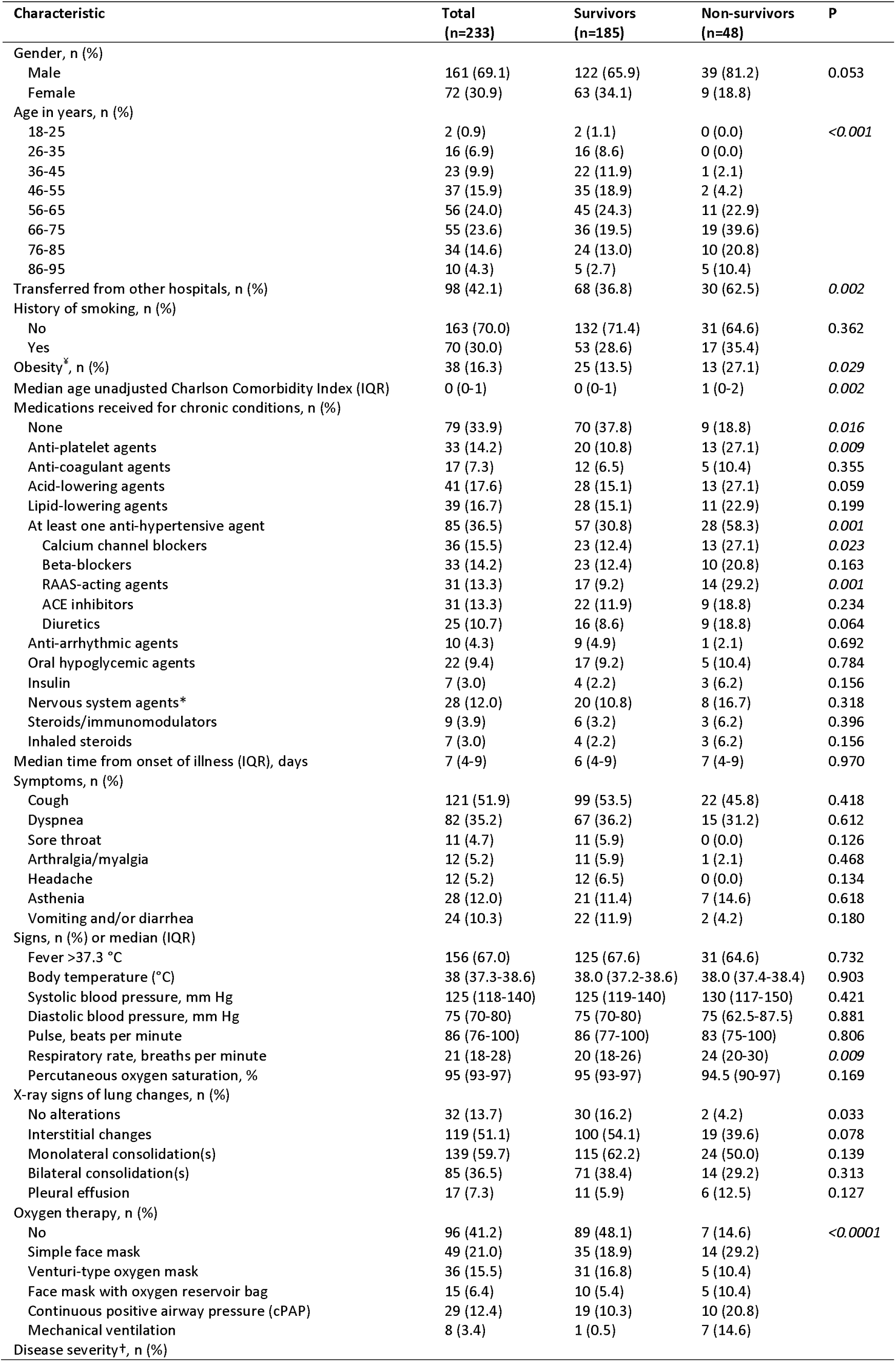
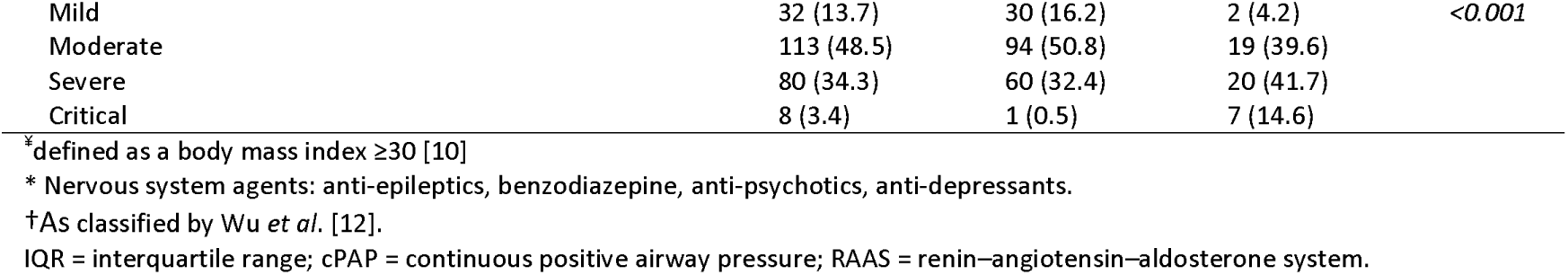
Characteristics upon hospital admission of the COVID-19 patients who or died during the study period

There were no significant between-group differences in terms of time from the onset of symptoms to hospital admission (overall median 7 days, IQR 4-9), and no significant differences in symptoms and signs upon admission. The most frequent presenting signs were fever, cough and dyspnea in both groups.

A higher proportion of survivors had chest X-rays without any pathological findings (16.2% *vs* 4.2%; p=0.033), but there was no difference in the pathological X-ray patterns between the two groups. A higher proportion of non-survivors presented with severe or critical disease (p<0.001).

Table 2 shows the baseline laboratory findings. The non-survivors had significantly lower median lymphocyte (p=0.008), hemoglobin (p=0.009) and albumin levels (p<0.001), and significantly higher median CRP (p<0.001), D-dimer (p<0.001) and creatinine levels (p<0.001).

**Table 2.** Laboratory findings upon admission

| Parameter | Median (IQR) or Number (%) |  |  | P |
| --- | --- | --- | --- | --- |
|  | Total<br>(n=233) | Survivors<br>(n=185) | Non-survivors<br>(n=48) |  |
| White blood cell count x 10<sup>9</sup>/L | 5.7 (4.4-7.5) | 5.5 (4.4-7.1) | 6.7 (4.8-9.0) | 0.006 |
| >10 x 10<sup>9</sup>/L | 24 (10.3) | 13 (7.0) | 11 (22.9) | 0.008 |
| Lymphocyte count x 10<sup>9</sup>/L | 1.0 (0.7-1.3) | 1.0 (0.8-1.4) | 0.9 (0.6-1.1) | 0.008 |
| <8.0 x 10<sup>9</sup>/L | 72 (30.9) | 52 (28.1) | 20 (41.7) | 0.081 |
| Neutrophil count x 10<sup>9</sup>/L | 4.1 (2.8-6.0) | 3.9 (2.8-5.3) | 5.7 (3.8-8.3) | <0.001 |
| Hemoglobin, g/dL | 13.8 (12.6-14.8) | 13.9 (13.0-14.8) | 13.2 (11.7-14.3) | 0.009 |
| Anemia* | 111 (47.6) | 79 (42.7) | 32 (66.7) | 0.003 |
| Platelets x 10<sup>9</sup>/L | 176 (137-221) | 174 (137-221) | 181 (141-226) | 0.534 |
| Prothrombin, INR | 1.2 (1.1-1.3) | 1.2 (1.1-1.3) | 1.2 (1.1-1.4) | 0.188 |
| Fibrinogen, g/L | 7 (6.3-7) | 7 (6-7) | 7 (7-7) | 0.148 |
| D-dimer, µg/L | 916 (535-1957) | 808 (494-1480) | 1740 (942-4851) | <0.001 |
| <500 | 51 (21.9) | 49 (26.5) | 2 (4.2) |  |
| 500-1000 | 77 (33.0) | 63 (34.1) | 14 (29.2) | 0.001 |
| > 1000 | 105 (45.1) | 73 (39.5) | 32 (66.7) |  |
| PaO<sub>2</sub>, mmHg | 69.5 (61-80.7) | 70.5 (62.7-81.2) | 64.5 (57.2-76.5) | 0.022 |
| C-reactive protein, mg/L | 47.6 (20.0-118.7) | 40.9 (17.9-102.0) | 130.0 (40.0-203.5) | <0.001 |
| < 50 | 124 (53.2) | 108 (58.4) | 16 (33.3) |  |
| 50-100 | 33 (14.2) | 28 (15.1) | 5 (10.4) | 0.001 |
| >100 | 76 (32.6) | 49 (26.5) | 27 (56.2) |  |
| Creatinine, mg/dL | 0.96 (0.79-1.2) | 0.93 (0.76-1.1) | 1.1 (0.96-1.68) | <0.001 |
| Lactate dehydrogenase, U/L (n= 229) | 338 (259-447) | 305 (246-404) | 438 (349-686) | <0.001 |
| Creatine kinase, U/L | 109 (62.0-237.0) | 93 (59.0-182.2) | 251 (106.7-392.0) | <0.001 |
| > 185 | 69 (29.6) | 43 (23.2) | 26 (54.2) | <0.001 |
| Alanine aminotransferase (U/L) | 32 (20-53) | 30 (20-49) | 40 (28-59) | 0.018 |
| Bilirubin, mg/dL | 1.2 (1.0-1.2) | 1.2 (1.1-1.2) | 1.1 (1.0-1.2) | 0.509 |
| Albumin, g/L (n=222) | 29 (25-33) | 30 (26-34) | 25 (23-30) | <0.001 |

\*Anemia defined as a hemoglobin value of <12.5 g/dL for females and <14 g/dL for males.
IQR = interquartile range; INR: International Normalised Ratio.

During the hospitalization 172 (73.8%) patients received a combination of LPV/r plus HCQ, 39 (16.7%) with remdesivir of whom 33 (14.2%) after LPV/r plus HCQ and 42 (18%) with tocilizumbab of whom 35 (83.3%) after LPV/r plus HCQ. Ten patients received all the 3 combinations. One hundred and forty-four (61.8%) patients were treated with at least one antibiotic during the hospital stay.

The median follow-up of the cohort as a whole was 40 days (IQR 33-47): eleven days (IQR 6-18) for the non-survivors and 44 days (IQR 38-49) for the survivors. The median hospital stay was 12 days (IQR 8-21). Kaplan Meier curve analysis showed that the time-dependent probability of death ten, 20 and 30 days after hospital admission was respectively 10.3% (95% confidence interval [CI] 6.4-14.2%), 16.3% (95% CI 11.6-21.1) and 19.7% (95% CI 14.6-24.9%) (Fig. 1).

**Figure 1.**
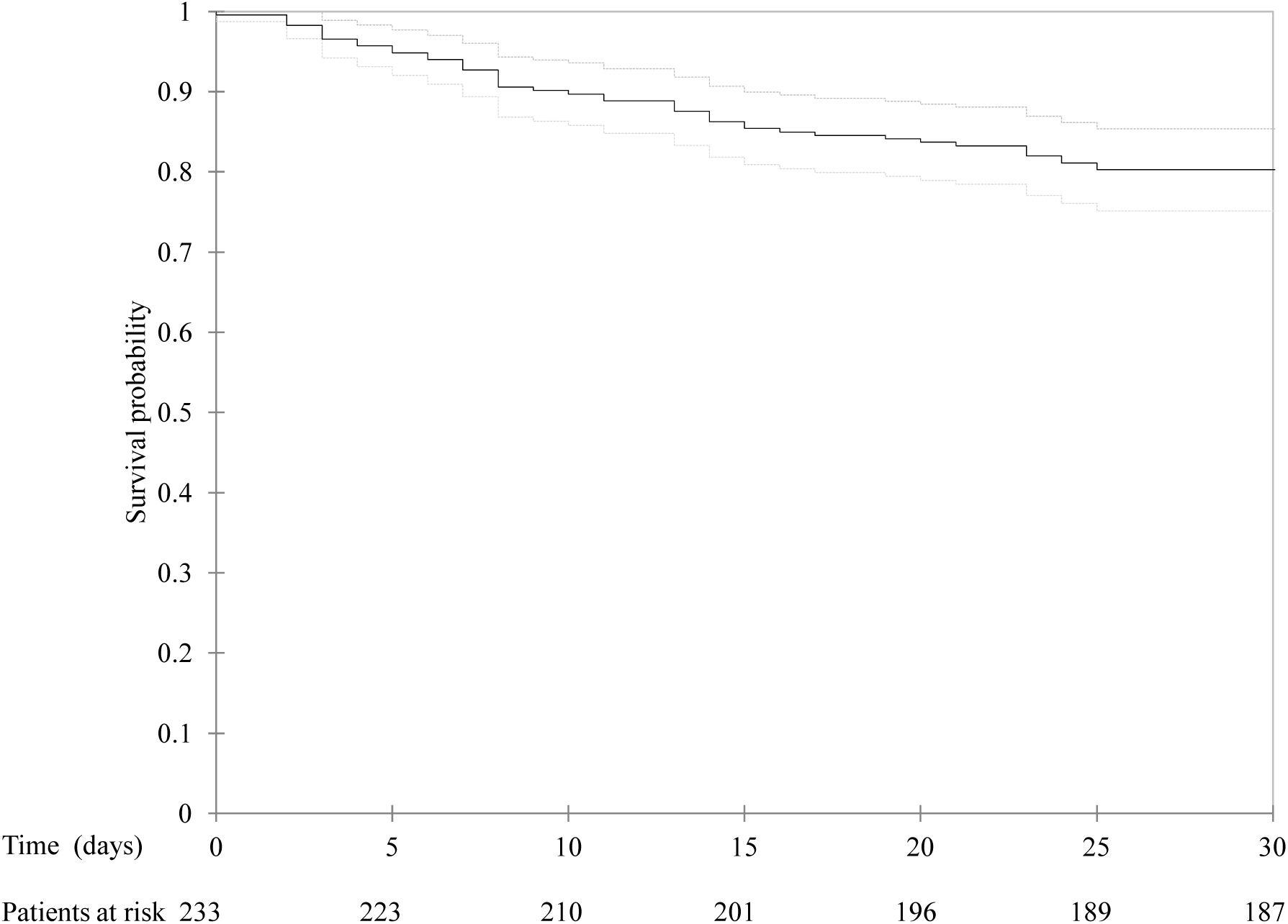
Kaplan-Meier curve of the probability of survival over time in patients with SARS-CoV-2 infection hospitalised in Milan, Italy. The continuous line represents the estimated survival curve, and the dashed lines the upper and lower limits of the 95% confidence interval.

Table 3 shows the uni- and multivariable Cox analysis of the factors associated with the risk of death. Age (adjusted hazard ratio [aHR] 2.08, 95% CI 1.48-2.92 per ten years more) and obesity (aHR 3.04, 95% CI 1.42-6.49) were independently associated with an increased risk of death, which was also associated with critical disease (aHR 8.26, 95% CI 1.41-48.29), CRP levels (aHR 1.17, 95% CI 1.02-1.35 per 50 mg/L more) and CK levels above 185 U/L (aHR 2.58, 95% CI 1.37-4.87) upon admission. Conversely, the multivariable model did not confirm the univariable findings of an increased risk of death in patients receiving at least one anti-hypertensive agent, those with anemia, or those with D-dimer levels of >1000 μg/L upon admission.

**Table 3.** Cox regression analysis of the demographic and clinical factors associated with SARS-CoV-2 infection mortality

| Characteristic | HR | 95% CI | P | aHR | 95% CI | P |
| --- | --- | --- | --- | --- | --- | --- |
| Gender |  |  |  |  |  |  |
| Female | 1 |  |  | 1 |  |  |
| Male | 2.02 | 0.98-4.16 | 0.058 | 1.42 | 0.62-3.28 | 0.409 |
| Age |  |  |  |  |  |  |
| Per 10 years more | 1.81 | 1.44-2.28 | <0.0001 | 2.08 | 1.48-2.92 | <0.0001 |
| Age unadjusted Charlson comorbidity index |  |  |  |  |  |  |
| Per one point more | 1.32 | 1.12-1.57 | 0.001 | 1.07 | 0.83-1.37 | 0.605 |
| Obesity<sup>*</sup> |  |  |  |  |  |  |
| No | 1 |  |  | 1 |  |  |
| Yes | 2.01 | 1.07-3.81 | 0.031 | 3.04 | 1.42-6.49 | 0.004 |
| Being treated with at least one anti-hypertensive agent |  |  |  |  |  |  |
| No | 1 |  |  | 1 |  |  |
| Yes | 2.78 | 1.56-4.94 | <0.001 | 1.41 | 0.73-2.74 | 0.309 |
| Disease severity<sup>†</sup> |  |  |  |  |  |  |
| Mild | 1 |  |  | 1 |  |  |
| Moderate | 2.82 | 0.66-12.10 | 0.163 | 1.30 | 0.29-5.77 | 0.727 |
| Severe | 4.43 | 1.04-18.97 | 0.045 | 1.50 | 0.32-7.04 | 0.605 |
| Critical | 35.35 | 7.29-171.43 | <0.0001 | 8.26 | 1.41-48.29 | 0.019 |
| Presence of anemia<sup>‡</sup> |  |  |  |  |  |  |
| No | 1 |  |  | 1 |  |  |
| Yes | 2.43 | 1.33-4.43 | 0.004 | 1.31 | 0.67-2.59 | 0.429 |
| Lymphocyte count |  |  |  |  |  |  |
| Per 100 cells/μL more | 0.91 | 0.00-0.97 | 0.008 | 0.98 | 0.91-1.06 | 0.664 |
| D-dimer |  |  |  |  |  |  |
| $\leq 1000$ μg/L | 1 | | | 1 | | |
| >1000 μg/L | 2.70 | 1.45-5.02 | 0.002 | 1.13 | 0.57-2.23 | 0.725 |
| C-reactive protein |  |  |  |  |  |  |
| Per 50 mg/L more | 1.26 | 1.15-1.38 | <0.0001 | 1.17 | 1.02-1.35 | 0.028 |
| Creatinine, |  |  |  |  |  |  |
| Per 0.5 mg/dL more | 1.08 | 1.01-1.15 | 0.030 | 1.0 | 0.92-1.1 | 0.528 |
| Creatine kinase |  |  |  |  |  |  |
| $\leq 185$ U/L | 1 | | | 1 | | |
| >185 U/L | 3.26 | 1.84-5.75 | <0.001 | 2.58 | 1.37-4.87 | 0.003 |

<sup>\*</sup> Obesity defined as body mass index $\geq 30$ points [10].
<sup>†</sup> Disease severity classification proposed by Wu *et al.* [12]: mild (mild clinical symptoms, no imaging signs of pneumonia); moderate (fever, cough, dyspnea or other symptoms, imaging signs of pneumonia); severe (any of: respiratory distress with respiratory rate (RR) of $\geq 30$ breaths per minute; resting oxygen saturation in air $\leq 93\%$ ; or $\text{PaO}_2 / \text{FiO}_2 \leq 300$ mmHg); and critical (any of: respiratory failure requiring mechanical ventilation; shock; or any other organ failure needing intensive care).
<sup>‡</sup> Anemia defined as hemoglobin $< 12.5$ g/dL for females, and $< 14$ g/dL for males.
HR = hazard ratio; aHR = adjusted hazard ratio; CI = confidence interval.

## DISCUSSION

We studied the characteristics and outcome of 233 adult COVID-19 patients hospitalised in Milan during the early dramatic days of the Italian epidemic. Forty-eight patients (20.6%) died during the study period, with the probability of dying at 30 days from hospital admission being of 19.7%. The overall case fatality rate observed in our cohort was similar to that found in a recent study of 201 COVID-19 patients hospitalised in Wuhan [13], but higher when compared to the 14% estimated by Wu *et al*. in the early period of the epidemic in China [14].

In line with the findings of some Chinese studies [13,15], our patients were prevalently male, which suggests a gender-based need for different hospital care during SARS-CoV-2 infection. It has been suggested that males may be more prone to developing severe and fatal COVID-19 [16, 17], and recent data regarding the epidemic in Europe shows a male-to-female death ratio of 2.1 that increases to 3.9 in patients aged 50-65 years [18]. However, our multivariable analysis did not reveal a significant gender-based difference in the survival rate. The male/female ratio among the patients admitted to our hospital during study period was 2.24, and so the reasons for the findings of the preliminary European report may be that females are less likely to be hospitalised with COVID-19.

The median age of our patients was 61 years which was similar to that reported in a recent large case series from the New York city area (63 years, IQR 52-75) [19], as against the 47-56 years reported in studies of Chinese hospitals [20, 21]; like the fact that the proportion of our patients aged >75 years was 18.9%, this difference probably mirrors the demographic differences between Italy and China [22]. Preliminary data from Chinese studies indicated that COVID-19 was more lethal in the elderly than younger people [13, 15], and we also found that an older age was an independent risk factor for death, as in the case of SARS and Middle East respiratory syndrome-related coronavirus [23, 24]. It has been speculated that older patients may be more likely to die of COVID-19 because age-related alterations in immunological functions and type 2 cytokine production lead to deficiencies in controlling SARS-CoV-2 replication and pro-inflammatory responses [15].

Our analysis showed that obese patients had a 3-fold higher risk of dying as compared to those with a body mass index below 30. This finding is in line with recently published studies suggesting that obesity represents one of the most important factors related to COVID-19 severity as evidenced by higher need of hospitalization and of invasive mechanical ventilation [25, 26]. Several mechanisms have been proposed to explain the increased severity of COVID-19 in obese patients, including the combination of reduced cardiorespiratory reserve and impairment of adaptive immune response to infections [27] It should be notice that the proportion of obese patients in our cohort was much lower than that recently described in a large case series by Richardson *et al* (16.3% *vs* 41.7%) and it cannot be excluded that in countries with high prevalence rate of obesity the effect of this condition on the burden of COVID-19 related mortality may be greater [19].

In line with the findings of previous Chinese studies [13, 15], fever, cough and dyspnoea were the most frequent signs/symptoms at the time of hospital admission; however, there was no significant difference between survivors and non-survivors in terms of symptoms upon admission.

The most frequent chest X-ray alterations were monolateral consolidations and interstitial alterations, followed by bilateral lung consolidations; however, no lung alterations were detected at x-ray in 13.7% of cases.

Most of our patients had mild or moderate disease [12] but, as expected, greater disease severity at the time of admission was strongly associated with an increased risk of death; in particular, the patients presenting with critical disease requiring assistance in ICU were at 8 times higher risk than those who did not.

In line with the study by Luo *et al* we found that serum CRP level upon admission was independently associated with adverse outcome of COVID-19 [28]. It has been shown that in pulmonary diseases marked by inflammatory features there is a typical raise in serum CRP level in response to inflammatory cytokines such as IL-6, IL-1 or TNF-α. [29]. Thus, higher CRP level in non-survivors of our study may indicate excessive and dangerous inflammatory response. We also found a significant correlation between higher CK levels upon admission and the risk of death. Elevated level of CK in COVID-19 patients might be a sign of respiratory muscle injury resulting from the increased demands placed on the respiratory system. There also previous evidence suggesting that serum levels of CK may rise in patients with pneumonia and pulmonary embolic disease [30]. Furthermore, it cannot be excluded that the increase in CK values in COVID-19 may be related to a damage to CK-rich tissues, such as skeletal and cardiac muscle and brain, directly induced by the virus or maladaptive immune responses. Autopsy studies are required to define the damage of organs that can cause CK increase during COVID-9.

Our study has a number of limitations. Firstly, as in China, the dramatically evolving scenario of the epidemic in Italy required continuous structural, organisational and staff changes that exposed the study to maturation bias. In particular, the number of laboratory examinations performed upon admission was limited (i.e. Interleukine-6 determination became available only after 15 days from the start of the study). Secondly, only 14.8% of our patients underwent a chest computed tomography upon admission because of barriers in our infrastructure created in order to ensure a dedicated COVID-19 service. Thirdly, it was difficult to ascertain the different effects on outcomes of the miscellaneous and often concomitant drug treatments given to our patients because of the absence of a standard of care for COVID-19 (excluding oxygen supplementation). Nevertheless, as all of the patients admitted to our Infectious Diseases Department were enrolled in this study and the study population was probably representative of the COVID-19 patients hospitalised in Italy in the early stage of the epidemic.

## CONCLUSIONS

Case-fatality rate of patients hospitalized with COVID-19 in the early days of the Italian epidemic was about 20%. Older age, obesity, disease severity upon admission were factors related with increased risk of death. Further studies are needed to evaluate pathogenic mechanisms of SARS-CoV-2 and the effect of the several proposed therapeutic approaches in reducing its lethality. Moreover, long term post discharge follow-up is warranted to provide a more accurate estimate of the morbidity and mortality attributable to this infection.

## Data Availability

The datasets used and/or analysed during the current study are available from the corresponding author on reasonable request.

## LIST OF ABBREVIATIONS

SARS-CoV-2: severe acute respiratory syndrome coronavirus 2; ICU: intensive care unit; CRP: C-reactive protein; CK: creatine kinase; cPAP: continuous positive airway pressure; RR: respiratory rate; LPV/r: lopinavir/ritonavir; HCQ: hydroxychloroquine; IQR: inter quartile range; CI: confidence interval; aHR: adjusted Hazard Ratio.

## CONSENT FOR PUBBLICATION

Not applicable.

## AUTHORS CONTRIBUTION

AG, ALR and MG designed the study. LO, AG were responsible for the statistical analysis. All authors contributed in the patient’s enrolment, data collection and interpretation. AG and ALR drawn a preliminary draft of the manuscript. MG, SA, SR, LM critically revised the manuscript. All authors approved the final version of the manuscript.

## AKNOWLEDGEMENTS

The authors would first like to thank all of the patients and all of the medical staff (paramedics, nurses and physicians) who begin this fight on one side of the wall and eventually fell ill during the battle. We would also like to thank Tiziana Formenti and Bianca Ghisi for their excellent and indefatigable technical help. Finally, our thanks go to all of the people who have given their unrestricted financial support by donating to our Institution.

